# Electrophysiological characteristics of lead-position-dependent EGM uninterrupted transition during left bundle branch pacing

**DOI:** 10.1101/2024.06.16.24308988

**Authors:** Jiabo Shen, Longfu Jiang, Hao Wu, Lu Zhang, Hengdong Li, Lifang Pan

## Abstract

**Background and Aims:** Left bundle branch pacing (LBBP) is a novel pacing strategy that improves ventricular synchrony by utilizing the native conduction system. However, the current standard practices limit continuous monitoring of paced electrocardiogram (ECG) and intracardiac electrogram (EGM) transition, which may result in overlooked or misinterpreted subtle transitions. This study aimed to explore the electrophysiological characteristics of the lead-position-dependent EGM continuous transition and evaluate their clinical significance.

**Methods:** This observational study included patients referred for LBBP due to symptomatic bradyarrhythmia. A continuous pacing and recording technique was employed, allowing real-time monitoring of progressive alterations in the paced QRS complex as the lead penetrates deeper into the ventricular septum. EGM and ECG parameters were continuously monitored and analyzed.

**Results:** The study encompassed 105 patients, with selective LBBP achieved in 88 patients (83.8%). The amplitude of ventricular EGM predictably changed with radial interventricular septum depth and peaked in the mid-septum. As the lead was inserted into the left ventricular subendocardium, the ventricular current of injury (COI) declined to a level approximating that of the right septum. Continuous recording technique enabled real-time monitoring of the entire perforation process and the subtle variations that exist among different perforation modalities. The discernment of discrete was feasible through the examination of unfiltered EGM, suggesting that selective LBB capture can also be confirmed by observing the subtle morphological transitions within the ventricular COI.

**Conclusions:** The continuous recording technique provides a more detailed understanding of the radial depth of the pacing lead throughout the implantation process. It simplifies the implantation procedures and facilitates the prevention or early detection of perforations. Future studies are needed to validate these findings and explore their clinical implications.

**What’s new?:**

1. Utilization of Ventricular Electrogram (EGM) for Lead Positioning: The amplitude of ventricular EGM changes predictably with radial interventricular septum depth, peaking in the mid-septum. This provides a useful way to determine whether the lead is located on the left, right, or middle of the ventricular septum.
2. Real-time Monitoring of Perforation Process: The continuous recording technique enables real-time monitoring of the entire perforation process. This feature helps to distinguish the subtle variations that exist among different perforation modalities, facilitating early detection and prevention of perforations.
3. Confirmation of Selective Left Bundle Branch Pacing (SLBBP): The emergence of a discrete ventricular current of injury (COI) may serve as a novel characteristic of SLBBP. This suggests that SLBBP can be confirmed by observing the subtle morphological transitions within the ventricular COI.

## Introduction

Left bundle branch pacing (LBBP), a novel pacing strategy, utilizes the native conduction system to improve ventricular synchrony, providing stable pacing parameters.[1] For the achievement of left conduction system capture, the LBBP lead must be embedded sufficiently deep within the left ventricular septum’s subendocardium.[2] Current standard practices of intermittent recording technique require the employment of an alligator clip for lead connection, a link disrupted during lead rotation, thereby inhibiting the capacity for continuous monitoring of paced QRS transition. Consequently, this may result in overlooked or misinterpreted subtle transitions. This limitation impedes the precise determination of left bundle branch (LBB) capture and any changes in the pacing modality as the lead advances into the ventricular septum.[3, 4] We have previously described a real-time recording technique that yields improved electrocardiogram (ECG) and intracardiac electrogram (EGM) recording capabilities in LBBP [5–7]. While previous studies have comprehensively described the lead-position-dependent QRS transition and output-dependent QRS and EGM transition,[7–10] the lead-position-dependent EGM transition remains inadequately established. Therefore, this study aims to explore the electrophysiological characteristics of the lead-position-dependent EGM continuous transition and evaluate their clinical significance.

## Methods

### Study population

This observational study encompassed consecutive patients referred to our center for LBBP, due to symptomatic bradyarrhythmia, from September 2022 through November 2023. Patients for whom definitive LBB capture could not be confirmed were excluded from the study. The research protocol was approved by the Institutional Review Board of Ningbo No.2 Hospital, and informed consent was duly obtained from each participant.

### Definitions of LBB capture

Dynamic paced QRS transition served as a criterion for LBB capture diagnosis.[7, 11, 12] LBB capture is confirmed by paced QRS morphology of the right bundle branch block pattern and all of the following strict criteria: (1) demonstration of the left ventricular septal pacing (LVSP) to nonselective LBBP (NSLBBP) transition during the process of lead screwing and/or NSLBBP to selective LBBP (SLBBP) transition during unipolar pacing threshold testing, and (2) both low and high outputs maintained the shortest and constant pacing stimulus to V6 R-wave peak time (Stim-V6RWPT).

### Implantation procedure

This study employs a continuous pacing and recording technique, utilizing a transseptal pacing lead as detailed in previous researches.[5, 7] Following a successful puncture of either the left axillary or subclavian vein, the C315 His sheath (Medtronic, Minneapolis, MN) is positioned in the right ventricle. Angiography is performed using 30° right anterior oblique fluoroscopy to visualize the tricuspid valve annulus (TVA), the image of which serves as a reference for the lead implantation site (**Figure 1A**).[13] Throughout the lead deployment process, a continuous pacemapping technique is adopted, establishing a persistent connection between the pacing lead and the pacing system analyzer (PSA) via the John Jiang connecting cable (Xinwell Medical Technology Co, Ningbo, Zhejiang, China).[5] In contrast to the interrupted pacing method, in which the alligator cable is momentarily disconnected during lead rotation, this strategy enables the real-time monitoring of progressive alterations in the paced QRS complex as the lead penetrates deeper into the interventricular septum (IVS).

**Figure 1.**
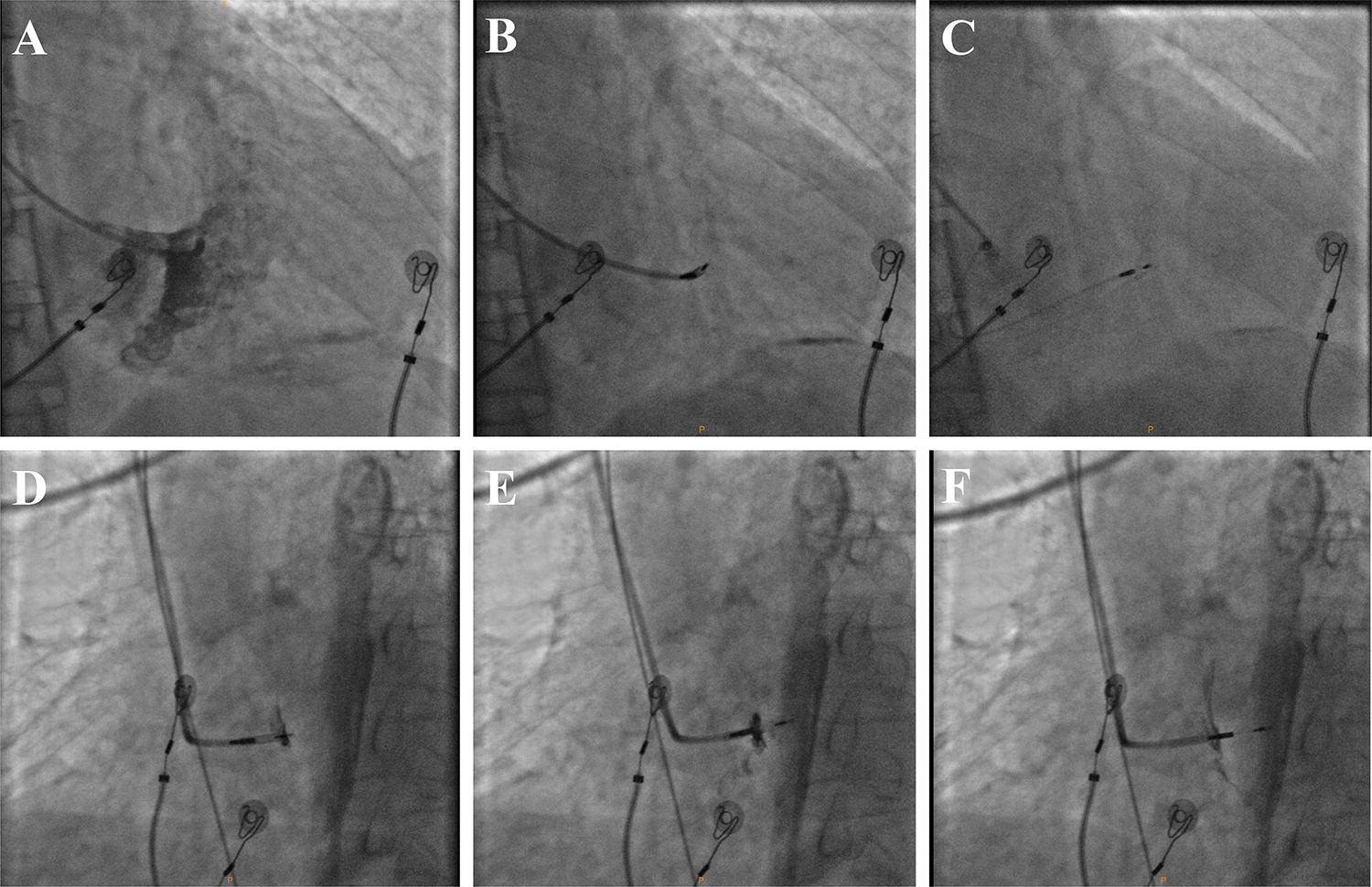
The procedure to implant the LBBP lead under fluoroscopy. A. Visualization of the TVA to help locate the LBBP target zone under the RAO 30° view. B-C. The 3830 lead was positioned in the LBBP screw-in site and fixed site under the RAO 30° view. D-F. Through the combination of septal angiography and the paced ECG and EGM morphologies associated, the RVSP, IVSP, and LBBP was confirmed. RAO, right anterior oblique; TVA, tricuspid valve annulus; RVSP, right ventricular septal pacing; IVSP, intraventricular septal pacing; LBBP, left bundle branch pacing; ECG, electrocardiogram; EGM, intracardiac electrogram.

The lead implantation procedure and schematic diagrams of the connection have been presented in earlier studies, illustrated through video documentation.[5, 7] The operational principle of the John Jiang connecting cable resembles a bearing, facilitating the free movement of internal and external components. This design ensures uninterrupted electrical conduction during lead rotation, thereby enabling continuous pacing and recording of ECG and EGM. The intricate internal structure and functional mechanism of the connector have been described in a previous study.[5] The distal pin of the 3830 lead (Medtronic, Minneapolis, MN) is inserted into the rotatable interface hole of the connector, after which the lead’s helix is inserted into the C315 His sheath and advanced into the right ventricle. Under fluoroscopic guidance, the 3830 lead is positioned at the LBB screw-in site, and the lead body is then rotated, gradually driving the helix from the right side of the ventricular septum to the left (**Figure 1B and C**). During this process, careful monitoring of changes in the ECG and EGM is crucial to accurately identify lead positioning and prevent perforation. The depth of the lead within the IVS is assessed by contrast injection from the sheath using a 45° left anterior oblique fluoroscopic view (**Figure 1D-F**).

### Electrophysiological observations

Continuous monitoring of ECG and EGM was conducted using the Workmate Claris (Abbott, Plymouth, MN) electrophysiology system at a speed of 100 mm/s. A filtered unipolar ventricular electrogram was obtained with high- and low-pass filter settings of 200 and 500 Hz to monitor the discrete EGM transition.[14] Unfiltered ventricular current of injury (COI) was monitored from the tip lead with band-pass filter settings of 0.5 and 500 Hz using a unipolar configuration during lead rotation.[15] The ventricular COI manifests as an increase in the duration and elevation of the ventricular electrogram compared to the baseline (**Figure 2**). Employing a continuous recording technique allows for the monitoring of changes in electrophysiological characteristics across all pacing modes during interventricular septum traversal. A detailed examination was conducted on five representative pacing modes within this study. Right ventricular septal pacing (RVSP) was defined when the lead was located in the RVS and exhibited the classic W pattern in lead V1. Intraventricular septal pacing (IVSP) was defined when the lead was screwed into the ventricular septum, and QRS morphological changes transitioned from the W pattern to the Qr pattern in lead V1. LVSP was defined when the lead was screwed into the deep septum, demonstrating a right bundle branch block pattern just before LBB capture. NSLBBP was defined when the Stim-V6RWPT for two adjacent paced complexes decreased by at least 10 ms with a constant output (2 V/0.5 ms) during screwing in. SLBBP was confirmed when the QRS morphology changed, the Stim-V6RWPT was fixed, and an isoelectric interval and discrete component appeared on the EGM (**Figure 3**). Measurements, including the maximum amplitude of ventricular COI elevation and other parameters, were performed by two individuals who were blinded to the study protocol. The values obtained from these measurements were subsequently averaged (**Figure 4**). In the event of identifying a perforation, characterized by a positive ventricular COI transition to a negative ventricular electrogram and high output loss capture, the lead was carefully withdrawn (**Figure 5**). During this withdrawal process, unipolar electrograms were continuously monitored to observe the recovery of positive ventricular COI. This observation served as a confirmation of the perforation.

**Figure 2.**
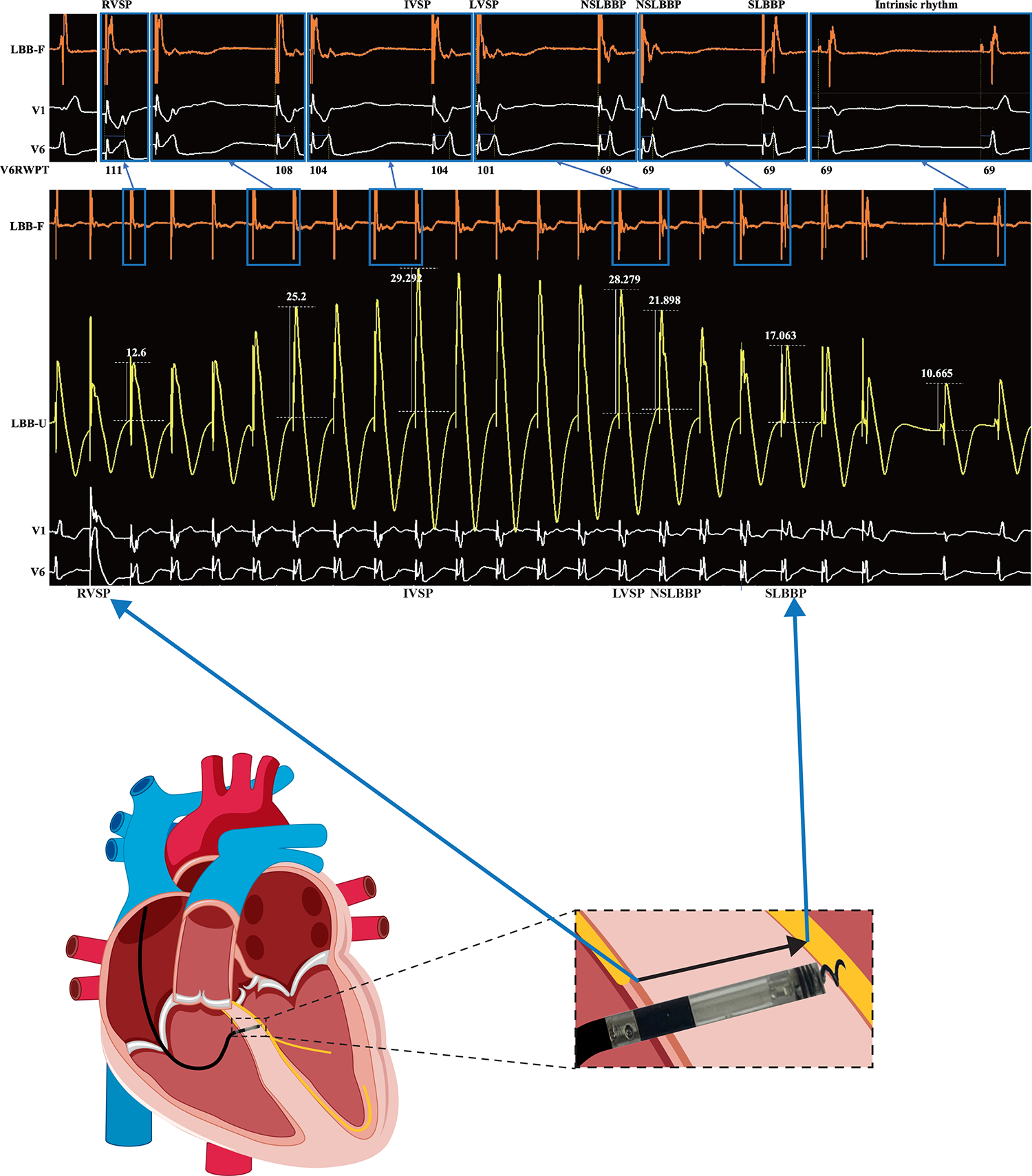
Lead-position-dependent EGM and ECG transition during screwing in. NSLBBP, nonselective left bundle branch pacing; SLBBP, selective left bundle branch pacing; LBB-F, filtered unipolar electrogram; LBB-U, unfiltered unipolar electrogram.

**Figure 3.**
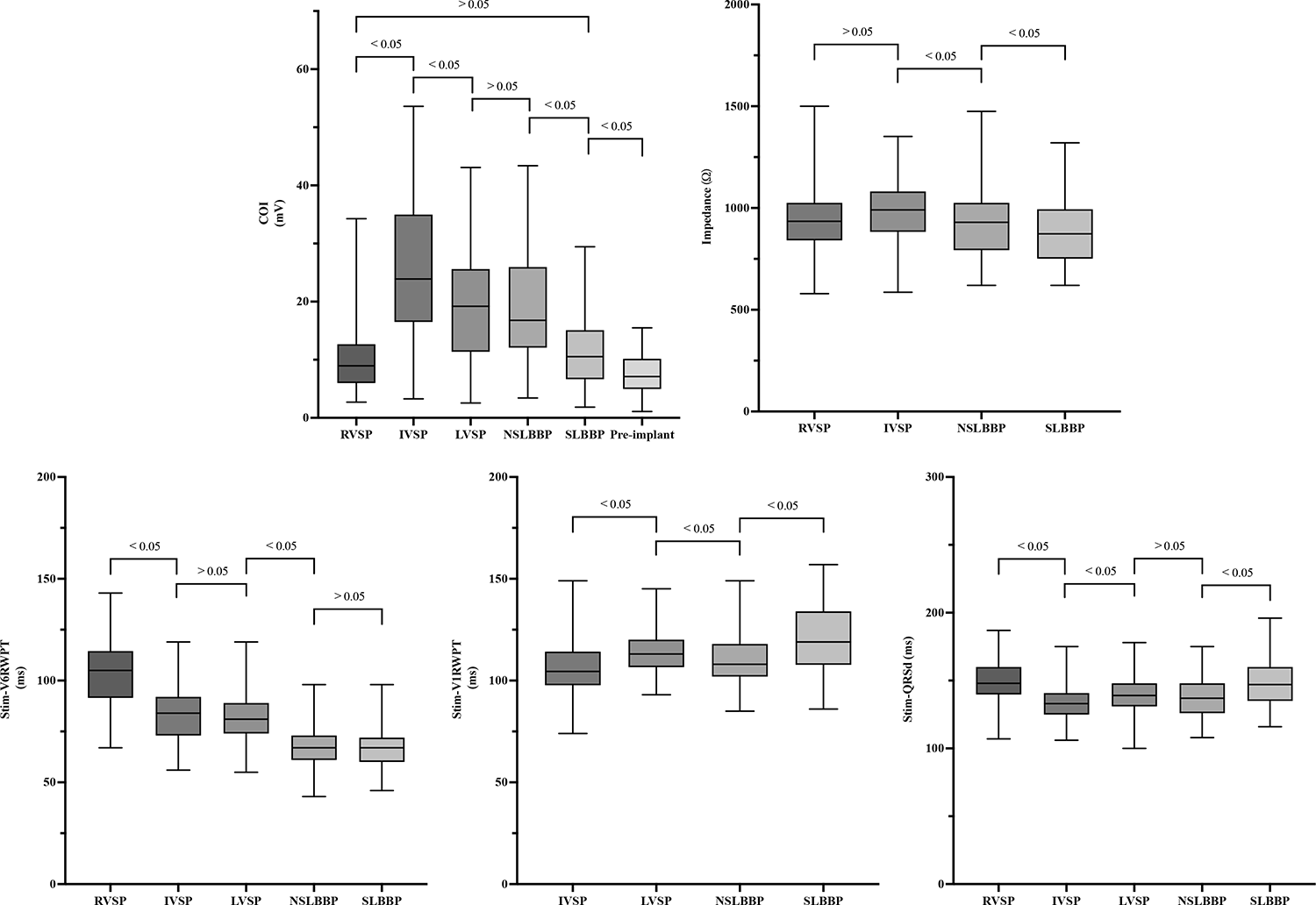
Electrophysiological characteristics of EGM and ECG in 5 pacing patterns. COI, current of injury; Stim-V6RWPT, stimulus to V6 R-wave peak time; Stim-V1RWPT, stimulus to V1 R-wave peak time. Abbreviations as in Figure 1 and 2.

**Figure 4.**
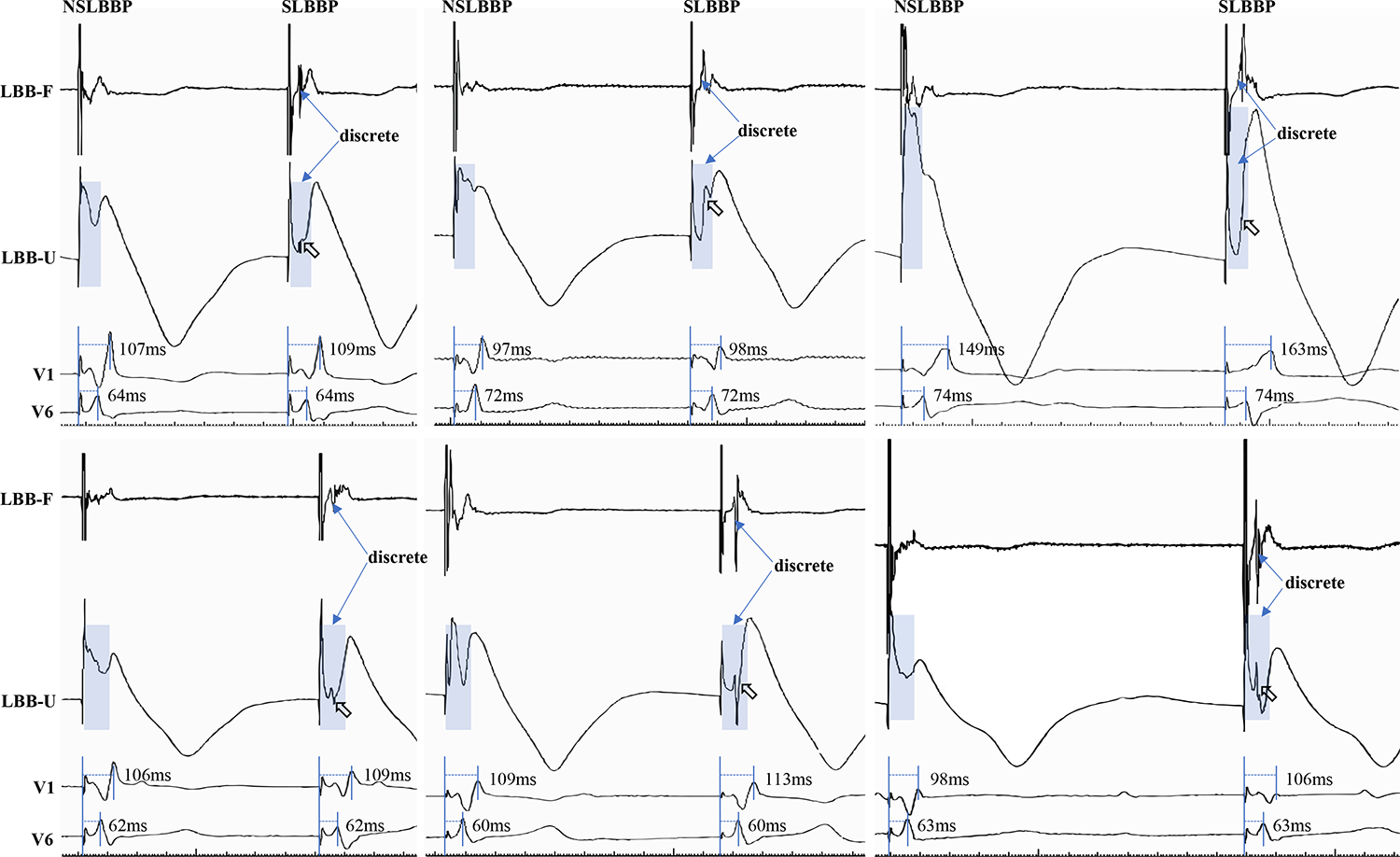
Discrete COI can be observed when the NSLBBP transition to SLBBP during threshold testing. Hollow arrows are used to indicate the appearance of notches. Abbreviations as in Figure 1 and 2.

**Figure 5.**
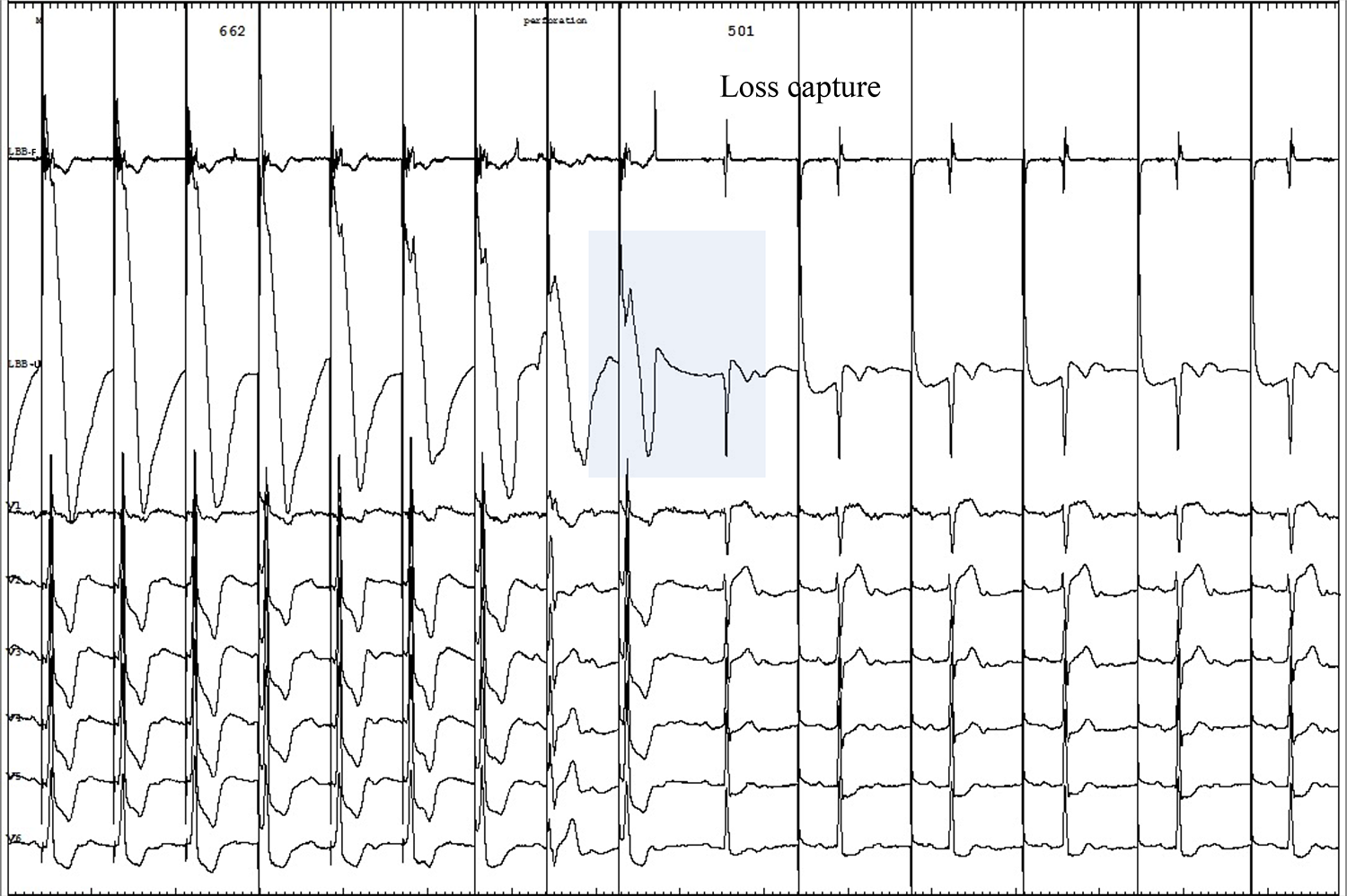
The continuous recording technique enables real-time monitoring of the entire perforation process, facilitating early detection of perforations. The impedance can be observed to drop from 662 Ω to 501 Ω. Abbreviations as in Figure 2.

### Statistical analysis

Continuous variables are represented as the mean ± standard deviation, while categorical variables are expressed in percentages. Statistical evaluation of differences among the ECG parameters within the six pacing modalities was conducted using repeated-measures analysis of variance complemented with Bonferroni post hoc analysis. A *p*-value of less than 0.05 was designated as the threshold for statistical significance. Statistical analyses were carried out utilizing SPSS (version 25.0; IBM Corp., Armonk, NY, USA) and GraphPad Prism 9 (GraphPad Software, Inc., San Diego, CA, USA).

## Results

### Baseline characteristics

This study encompassed a total of 105 patients who underwent pacemaker implantation. NSLBBP was confirmed in ninety-four patients (89.5%) who displayed an abrupt shortening of Stim-V6RWPT by at least 10 ms. SLBBP was achieved in 88 patients (83.8%), as indicated by isoelectric intervals and discrete components during threshold testing. In the absence of a Stim-V6RWPT’s abrupt shortening of 10 ms and an isoelectric interval, LVSP was confirmed in eleven patients.

An analysis was performed on paced parameter changes in the 88 patients who met the criteria for selective LBB (SLBB) capture. The mean age of the patients was 73.7 ± 9.1 years. The left ventricular ejection fraction was reported as 62.8 ± 11.1%. The indications for pacemaker implantation included atrioventricular block in 56 patients and sick sinus syndrome in 31 patients. In the pacemaker population, hypertension was identified in 53 patients, diabetes mellitus in 23 patients, cardiomyopathy in 4 patients, coronary heart disease in 14 patients, and atrial fibrillation in 22 patients. **Table 1** provides a summary of the patients’ characteristics. The pacing-related and procedure-related characteristics are presented in **Table 2**.

**Table 1.** Patient characteristics (n = 88)

|  |  |
| --- | --- |
| Age (years) | 73.7 ± 9.1 |
| Male | 51 (58.0) |
| Pacing indication, n (%) |  |
| Atrioventricular block | 56 (63.6) |
| Sick sinus syndrome | 31 (35.2) |
| Atrial fibrillation with bradycardia | 4 (4.5) |
| Heart failure | 5 (5.7) |
| Comorbidities, n (%) |  |
| Hypertension | 53 (60.2) |
| Diabetes mellitus | 23 (26.1) |
| Cardiomyopathy | 4 (4.5) |
| Coronary heart disease | 14 (16.0) |
| Atrial fibrillation | 22 (25.0) |
| LVEF (%) | 62.8 ± 11.1 |
| LVDD (mm) | 49.7 ± 7.8 |
| QRS morphology, n (%) |  |
| Narrow QRS | 60 (68.2) |
| RBBB | 19 (21.6) |
| LBBB | 11 (12.5) |
| NIVCD | 2 (2.3) |

Values are given as mean ± SD or n (%).
LVEF, left ventricular ejection fraction; LVDD, left ventricular end-diastolic dimension; RBBB, right bundle branch block; LBBB, left bundle branch block; NIVCD, non-specific intraventricular conduction disturbance.

**Table 2.**
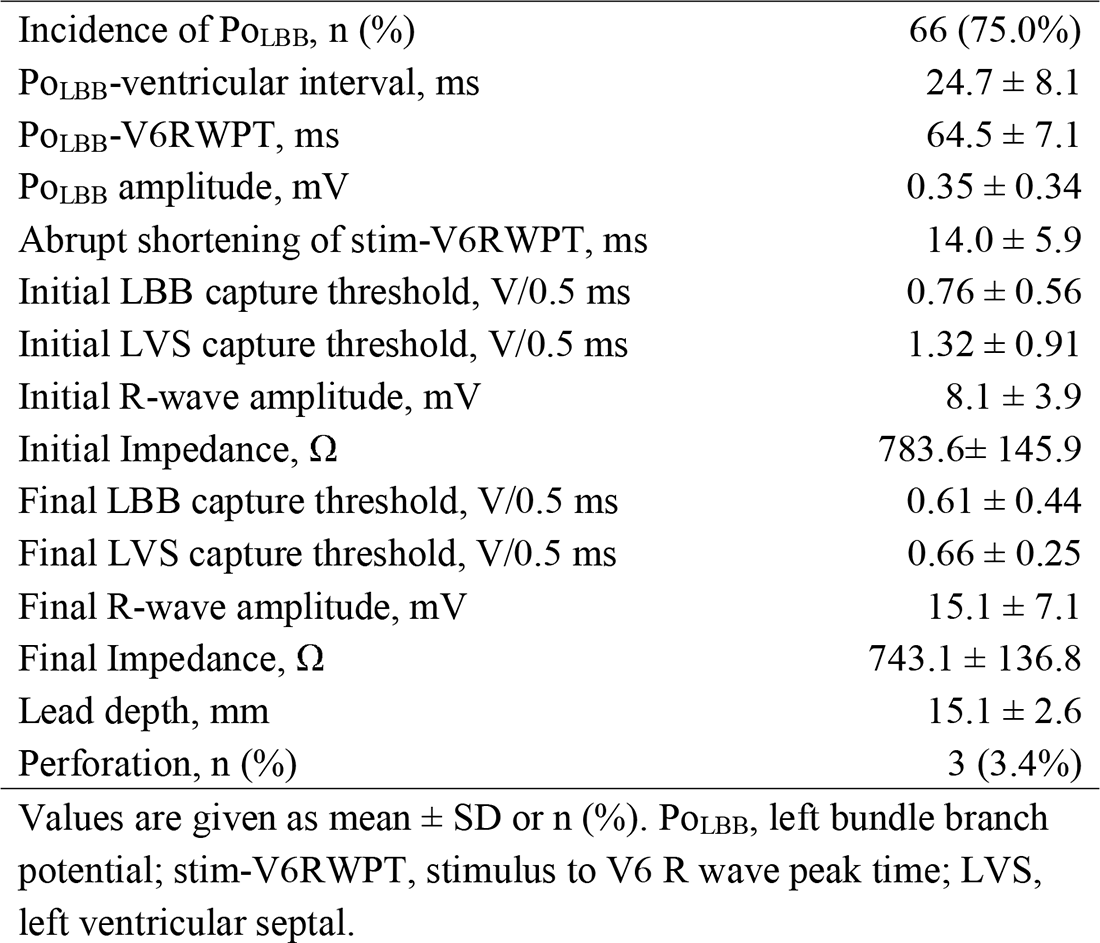
Procedure-related parameters (n=88)

### Lead-position-dependent parameters during implantation

The ECG and EGM parameters at the depth of implantation are documented in **Table 3** and **Figure 3**. The ventricular COI amplitude was recorded as 10.5 ± 5.9 mV in the RVSP group and 11.9 ± 6.4 mV in the SLBBP group, with no significant difference (*p* = 0.328). There was no significant difference in the ventricular COI amplitude between the LVSP and NSLBBP groups (19.8 ± 9.6 mV vs. 20.0 ± 9.2 mV). A significant statistical difference was observed in the amplitude of ventricular COI among other pacing modes at varying depths.

**Table 3.** Lead-position–dependent parameters during LBBP.

|  | RVSP | IVSP | LVSP | NSLBBP | SLBBP | Pre-pac<br>impla |
| --- | --- | --- | --- | --- | --- | --- |
| COI (mV) | 10.5±5.9 | 26.6±11.8 | 19.8±9.6 | 20.0±9.2 | 11.9±6.4 |  |
| Impedance ($\Omega$) | 948.7±202.1 | 980.3±156.3 | NA | 929.5±191.4 | 886.0±164.4 | 743.1 |
| Stim-V6RWPT (ms) | 103.2±16.9 | 83.8±12.5 | 81.8±12.3 | 67.5±9.7 | 66.9±9.5 | 70. |
| Stim-V1RWPT (ms) | NA | 105.8±12.0 | 113.7±11.0 | 110.4±12.1 | 118.4±16.1 | 114. |
| Stim-QRSend (ms) | 148.0±14.9 | 133.9±13.8 | 138.6±14.3 | 136.0±18.8 | 147.5±17.6 | 144. |

RVSP, right ventricular septal pacing; IVSP, intraventricular septal pacing; LVSP, left ventricular septal endocardium pacing; NSLBBP, non-selective left bundle branch pacing; SLBBP, selective left bundle branch pacing; COI, current of injury; RWPT, R wave peak time; Stim-QRSend, stimulus-QRS end duration.

Impedance was measured as 948.7 ± 202.1Ω in the RVSP group and 980.3 ± 156.3 Ω in the IVSP group, with no significant difference (*p* = 0.555). A significant difference in impedance was noted between the IVSP group (980.3 ± 156.3 Ω) and the NSLBBP group (929.5 ± 191.4 Ω) (*p* < 0.05), as well as between the NSLBBP group (929.5 ± 191.4 Ω) and the SLBBP group (886.0 ± 164.4 Ω) (*p* < 0.05).

Stim-V6RWPT was 103.2±16.9 ms in the RVSP group and 83.8 ± 12.5 ms in the IVSP group, with a significant difference (*p* < 0.05). There was no significant difference in Stim-V6RWPT between the IVSP and LVSP groups (83.8 ±12.5 ms vs. 81.8±12.3 ms). However, Stim-V6RWPT significantly shortened from LVSP to NSLBBP (67.5 ± 9.7 ms, *p* < 0.05), with no significant difference between the NSLBBP and SLBBP groups (66.9 ± 9.5 ms).

Significant statistical differences were observed in stimulus to V1 R-wave peak time with different depth pacing modes. No significant difference was noted in stimulus-QRS end duration (Stim-QRSend) between the LVSP and NSLBBP groups (138.6 ± 14.3ms vs. 136.0 ± 18.8 ms). However, a significant difference in Stim-QRSend was observed between the NSLBBP group (136.0 ± 18.8 ms) and the SLBBP group (147.5 ± 17.6 ms) (*p* < 0.05).

Apart from LVS perforation experienced by three patients, no other procedure-related complications, such as electrode displacement, pericardial effusion, infection, or pocket hematoma, were reported.

## Discussion

Our study demonstrates several novel findings: 1) The amplitude of ventricular EGM predictably changes with radial IVS depth, and peaks in the mid-interventricular septum, making it a useful ventricular COI feature to distinguish whether the lead is located on the left, right, or middle of the ventricular septum; 2) As the lead is inserted into the left ventricular subendocardium, producing a decline in the ventricular COI to a level approximating that of the right septum, the operation necessitates an extraordinarily cautious screwing of the lead. Simultaneously, careful observation of the QRS transition is necessitated along with the execution of output testing; 3) Continuous recording technique enables real-time monitoring of the entire perforation process. This feature helps to distinguish the subtle variations that exist among different perforation modalities; 4) The discernment of discrete is feasible through the examination of unfiltered EGM. This suggests that SLBB capture can also be confirmed by meticulously observing the subtle morphological transitions within the ventricular COI.

### Mid-interventricular septal pacing

During the implantation procedure of cardiac devices, acute tissue injury, also known as the COI, can be caused by the fixation of leads into the myocardial tissue. The presence of COI has been associated with improved acute performance and long-term stability of active fixation leads.[16–18] It is generally accepted that a deeper electrode screw-in leads to increased tissue damage, consequently resulting in higher COI amplitudes. Intriguingly, our study reveals that while deeper electrode insertion may potentially cause increased myocardial damage, it is not positively correlated with COI elevation. Instead, it exhibits an inverted U-shaped relationship, resembling a parabolic curve. This suggests that the injury current might be associated not solely with myocardial damage, but also with the depth at which the lead is positioned within the ventricular septum. If the COI does not increase when the electrode is initially inserted under the right septal imaging positioning, it implies that the electrode is entangled with the endometrium or myocardial fibrosis, necessitating a change in position. The peak of the ventricular COI is not observed in the left deep septum; rather, it is in the mid-interventricular septum where the maximum amplitude is recorded. Positioning of the lead in the mid-interventricular septum may suggest its optimal stability.

Evidence from previous studies indicates that IVSP is characterized by a relatively short RWPT and QRS duration, indicative of a superior level of synchronicity between the left and right ventricles associated with this pacing modality. Our research also suggests the potential for this modality to offer superior stability. Consequently, given the current lack of consensus regarding the optimal strategy for trans-septal pacing, IVSP may be considered a viable choice for practitioners seeking to attain the greatest lead stability and good ventricular synchrony. Compared to left-side deep septal pacing (including LVSP and LBBP), the application of IVSP may be more straightforward and presents a lower risk of perforation.

### Deep septal pacing lead localization and perforation

The implantation technique of LBBP involves inserting a lead into the deep septum below the left ventricular subendocardium to engage the LBB. The number of rotations required to access the LBB area is dependent on factors such as septal thickness, the degree of fibrosis, and sheath support. The assessment of ventricular perforation can be conducted using X-ray fluoroscopy and echocardiography, but the precise positioning of the intramyocardial lead often remains unclear due to the lack of soft tissue visualization in X-ray fluoroscopy and off-axis imaging planes in echocardiography. Typically, lead position is determined through the observation of ECG changes, while perforation detection relies on monitoring impedance changes and threshold assessments. However, impedance values can be significantly influenced by the quality of tissue-electrode contact, which varies during rapid screwing, making it challenging to accurately respond to real-time impedance changes.[19] Consequently, delays might occur in identifying perforation via impedance and threshold evaluations.

Septal perforation could be associated with the operator’s endeavor to achieve the best possible paced QRS morphology and the LBB capture. This highlights the clinical need for the precise localization of the lead. The ventricular COI, represented by the unfiltered intracardiac electrical signal detected at the lead’s tip, can accurately measure the contact between the lead and myocardial tissue, suggesting its potential to subtly discern ventricular perforation. This sensitivity primarily arises since any disruption in the lead-myocardium contact is immediately mirrored in this measurement. However, the current intermittent recording technique, which requires disconnection during lead screwing, limits the widespread use of this indicator by eliminating the possibility of real-time tracking of dynamic intracardiac electrical signal changes. Consequently, it becomes impossible to definitively confirm the occurrence of perforation during this process. Confirmation can only be achieved once reconnection between the pacing lead and the external PSA is established, undeniably prolonging the procedure and increasing the risk of complications.

The continuous recording technique addresses these limitations. As demonstrated in our research, the amplitude of the ventricular COI allows for real-time determination of the radial depth of the lead within the interventricular septum, thereby facilitating the prevention and detection of perforations. During the entire screw-in process, it was observed that the amplitude of the ventricular COI increases and peaks as the lead is inserted from the right septum to the mid-septum. As the lead continues to be inserted deeper into the left septum, the ventricular COI amplitude gradually decreases, eventually approximating the amplitude initially detected within the right septum. In our study, it was observed that the ventricular COI from both the right and left septum were closely aligned. Consequently, it was inferred that by monitoring the COI of the right septum, the ventricular COI amplitude of the left septum could be predicted. This insight enabled us to determine, with greater precision, the individualized endpoint for halting the screwing into the left ventricular endocardium, based on the COI amplitude in the right ventricular septum. At this stage, the lead should be manipulated with care to prevent a sudden drop in the ventricular COI. Simultaneously, dynamic changes in the ECG, such as an abrupt shortening of V6RWPT and the emergence of an initial r wave in the V1 lead, need to be closely monitored. Once these changes are detected, the rotation of the lead should be immediately terminated and threshold testing conducted, typically revealing a discrete ventricular electrogram.

Even in the face of complications such as ventricular perforation, the advantages of the continuous recording technique allow for the rapid detection of a sudden decrease in ventricular COI, enabling early termination of lead rotation and a change in the implantation location. Simultaneously, this process discerns the nature of the perforation. It enables differentiation between a micro-perforation, where the helix partially enters the left ventricle cavity, and a complete perforation, where the helix fully penetrates the ventricular septum. With micro-perforations, the ventricular COI can still be observed, whereas complete perforations typically present as a QS pattern.[20]

### Selective left bundle branch pacing

The gold standard for confirming LBB capture typically involves observing a transition in QRS morphology, a change that indicates differences in excitability (as measured by threshold testing) and/or refractoriness (as determined by programmed stimulation) between the LBB and myocardium.[11] Wu et al. demonstrated that an LBB potential can still be recorded by the lead even when positioned a certain distance away from the LBB, suggesting that the presence of an LBB potential does not necessarily confirm LBB capture.[21] A previous study proposed that the LBB COI reflects the anatomical location of the tip of the lead within the LBB region, possibly due to localized cell membrane damage caused by the pressure exerted by the electrode on the LBB. Therefore, recording an LBB potential with COI can serve as an indicator of LBB capture.[22] However, this method is only applicable to patients without left bundle branch block, as a considerable proportion of patients do not have Po_LBB_ and thereby LBB COI, and its applicability is somewhat limited.

As previous studies have shown, the presence of ventricular COI is associated with adequacy of lead fixation in the endocardium.[16–18] It is generally believed that ventricular COI itself is not diagnostic for LBB capture.[23] In our study, besides finding that the characteristics of COI vary at different septum depths, small and significant changes in ventricular COI can also be observed when the NSLBB transition to SLBB capture. Just as filtered electrograms can display discrete EGM, unfiltered electrograms can also display discrete ventricular COI. Previous research has shown that the physiological Purkinje activation was like distal to proximal activation of the ventricular component.[24] A discrete EGM in the pacing lead can be recorded because direct myocardial capture is absent and therefore ventricular activation over the pacing lead occurs late following initial conduction only over the LBB-Purkinje system. Recording discrete EGM was defined as SLBBP and had a specificity of 100% for confirmation of LBB capture, which was demonstrated by a previous study.[25] Therefore, the discrete ventricular COI can also serve as an electrophysiological phenomenon for capturing the LBB.

## Conclusion

In this study, we present a detailed description of the electrophysiological properties of unipolar lead-position-dependent EGM. Such insights offer clinicians a subtle understanding of the radial depth of the pacing lead throughout the implantation process. Given that IVSP results in the most favorable ventricular COI, this strategy provides operators with a more selection of pacing locations. Furthermore, the emergence of a discrete ventricular COI may serve as a novel characteristic of SLBBP. Consequently, the use of continuous recording technique enables the observation of a broader array of electrophysiological phenomena during traversal of the IVS. This approach not only provides practitioners with more insights but also simplifies implantation procedures and facilitates the prevention or early detection of perforations. Future studies are needed to further validate these findings and explore their clinical implications.

### Study Limitations

This study has several limitations that should be noted. Firstly, the study was conducted at a single center, and the sample size was relatively small. This may limit the generalizability of our findings to broader populations or different clinical settings. Secondly, the observational nature of the study design precludes the determination of causality. Future studies with randomized controlled trials are needed to establish causal relationships between the variables we studied. Thirdly, we used specific equipment and techniques for the continuous recording of electrophysiological phenomena during left bundle branch pacing. The results might not be applicable to centers using different types of equipment or techniques. Lastly, although our study provides evidence that the continuous recording technique can facilitate the observation of a broader array of electrophysiological phenomena during traversal of the IVS, more research is needed to determine whether this approach can improve clinical outcomes, such as reducing the incidence of complications or improving the long-term stability of the pacing lead.

## Data Availability

All data produced in the present study are available upon reasonable request to the authors

